# Cognitive and Cholinergic Systems Trajectories in Parkinson Disease

**DOI:** 10.1101/2024.07.17.24310588

**Authors:** Taylor Brown, Prabesh Kanel, Giulia Carli, Jaimie Barr, Nicolaas I. Bohnen, Roger L. Albin

## Abstract

**Objective:** Cognitive decline in Parkinson disease (PD) is a disabling and highly variable non-motor feature. While cholinergic systems degeneration is linked to cognitive impairments in PD, most prior research reported cross-sectional associations. We aimed to fill this gap by investigating whether baseline regional cerebral vesicular acetylcholine transporter ligand [^18^F]-fluoroethoxybenzovesamicol ([^18^F]-FEOBV) binding predicts longitudinal cognitive changes in mild to moderate, non-demented PD subjects.

**Methods:** Seventy-five non-demented, mild-moderate PD subjects received baseline standardized cognitive evaluations and [^18^F]-FEOBV PET imaging with repeat cognitive evaluations 2 years later. Participants were classified into four cognitive classes based on stability or change in cognition: Persistent normal (no MCI at baseline and follow-up), Persistent MCI, MCI conversion, and MCI reversion. Whole-brain voxel comparisons with normal controls, and voxel-based and cluster volume-of-interest correlation analyses with longitudinal cognitive changes were performed.

**Results:** Whole-brain voxel comparisons of each class with a matched control group revealed unique bi-directional differences in baseline regional [^18^F]-FEOBV binding. Increased regional [^18^F]-FEOBV binding in predominantly anterior cortical and sub-cortical regions was found in the persistent normal and MCI reversion groups. Whole-brain voxel correlation analysis between baseline [^18^F]-FEOBV binding and two-year longitudinal percent changes in cognition identified a specific regional pattern of reduced posterior cortical, limbic and paralimbic [^18^F]-FEOBV binding predictive of global cognitive declines and across five cognitive domains at two-year follow-ups.

**Interpretation:** Cholinergic system changes correlate with varying cognitive trajectories in mild-moderate PD. Upregulation of cholinergic neurotransmission may be an important compensatory process in mild-moderate PD.

## INTRODUCTION

Cognitive impairments are debilitating non-motor complications of Parkinson disease (PD) associated with substantial disability and impaired quality of life.^1^ Approximately 20-30% of PD patients exhibit Mild Cognitive Impairment (MCI) at diagnosis, with up to 80% developing dementia over a 20-year follow-up period.^2^ Cognitive decline trajectories vary widely among individual PD patients, with some progressing to dementia within a few years after diagnosis, others exhibiting slow progression or fluctuating cognitive function, and still others maintaining intact cognition for extended periods.^3–5^ Despite the shared presence of striatal dopaminergic denervation, impairments across different cognitive domains and degrees of involvement in progression of cognitive impairments are variable.^3,6,7^ Heterogeneity in rates of progression of cognitive deficits and variable domain involvements suggests that pathologies and mechanisms outside the dopaminergic system contribute to cognitive deficits in PD.

Post-mortem neurochemical and anatomic studies identified individuals with Parkinson disease dementia (PDD) as exhibiting marked cholinergic deficits in the basal forebrain corticopetal cholinergic system.^8,9^ Advancements in imaging techniques subsequently allowed *in vivo* assessments of regional cholinergic synapse and nerve terminal densities with the majority of studies utilizing acetylcholinesterase (AChase; [^11^C]-PMP, [^11^C]-MP4) substrate PET or vesicular acetylcholine transporter (VAChT; [^18^F]-FEOBV) ligand PET.^10,11^ Cross-sectional imaging studies confirmed post-mortem research associating cholinergic deficits with cognitive decline. In PD participants with and without dementia, diffuse low cortical acetylcholinesterase activity correlates robustly with cognitive impairments.^12,13^ Other cross-sectional studies identified distinct cortical cholinergic deficit topographies associated with cognitive impairments in PD.^12,14,15^ These studies predominantly identified posterior cortical cholinergic deficit correlates of cognitive impairments in PD.^12,16^ While most research focused on cholinergic denervation in PD and correlates with clinical features, bidirectional alterations of [^18^F]-FEOBV binding or AChase activity were demonstrated in some studies.^17,18,19^ Van der Zee et al. demonstrated significantly higher [^18^F]-FEOBV binding in *de novo* PD with normal cognition and PD with MCI compared to HC, identifying increased [^18^F]-FEOBV binding within orbito-frontal, cerebellar, and anterior cingulate regions in a normal cognition PD group.^17^ These findings suggest cholinergic system change correlates of cognitive status in PD with potential compensatory cholinergic mechanisms in both cognitively normal and cognitively impaired PD.^17,18,19^ The majority of cholinergic system PET research on cognitive changes in PD was cross-sectional.^14,17–21^ A recent longitudinal study utilizing AChase PET indicated complex patterns of progression of regional cholinergic synapse vulnerability with evidence of posterior-anterior progression of cholinergic deficits in frontal cortices.^21^

Given robust cross-sectional correlates, additional longitudinal research is crucial to explore the contribution of cholinergic systems changes in cognitive decline in PD and for determining potential predictive use of cholinergic systems changes for cognitive declines in PD. Building on prior cross-sectional studies, we aimed to determine whether baseline [^18^F]-FEOBV imaging in a cognitively heterogeneous group of PD patients predicts cognitive changes over a two-year period and explore potential differences in regional cholinergic terminal deficits between subgroups with different cognitive trajectories.

## MATERIALS AND METHODS

### Ethical Compliance

Written informed consent was obtained from all participants according to the Declaration of Helsinki. This study was approved by the Institutional Review Boards of the University of Michigan School of Medicine and Veterans Affairs Ann Arbor Healthcare System. This study was registered at ClinicalTrials.gov (NCT02458430 & NCT01754168).

### Participants

PD subjects were recruited from the Movement Disorders Clinics at the University of Michigan and affiliated VA Ann Arbor Health System. PD participants met the UK Parkinson’s Disease Society Brain Bank Clinical diagnostic criteria. Typical striatal dopaminergic denervation was confirmed in all PD participants by [^11^C]-dihydrotetrabenazine PET. Potential subjects with evidence of large vessel stroke or other intracranial lesions on anatomic imaging were excluded. No enrolled participant was demented, using anti-cholinergic agents, or cholinesterase inhibitor drugs. Two participants were actively using tobacco products. A healthy control (HC) group consisting of 17 men and 19 women with mean age 68.03 (SD 6.49) years was included for normative [^18^F]-FEOBV PET imaging data. HC subjects did not have a history of neurological or psychiatric disorders.

### Clinical Assessments

PD participants underwent standardized assessments at baseline and 2-year follow-up visits. Assessments included demographic data, medication history, Instrumental Activities of Daily Living, Montreal Cognitive Assessment, Movement Disorder Society-Unified Parkinson Disease Rating Scale evaluations in the overnight “OFF” state, and a comprehensive neuropsychological test battery examining five cognitive domains: attention, executive, language, memory, and visual (one visuospatial and one visuo-constructive assessment) (**Supplementary Table 1**). Each cognitive domain evaluation consisted of at least two sub-tests from the comprehensive neuropsychological test battery (**Supplementary Table 1**).^19^ To standardize results and assess longitudinal changes, cognitive testing results were expressed as z-scores. Z-scores were calculated based on a dataset collected in this laboratory of older participants of similar ages, sex, and educational levels. For each cognitive domain, domain z-scores were computed by averaging z-scores of relevant sub-tests. Global cognition composite z-scores were then computed by averaging the z-scores of each cognitive domain. Domain specific and global cognitive composite z-scores were determined at baseline and again for the two-year follow-ups.

### Imaging acquisition and pre-processing

Brain MRI was performed on a 3 Tesla Philips Achieva system (Philips, Best, The Netherlands) and PET imaging was performed in 3D imaging mode with a Biograph 6 TruPoint PET/CT scanner (Siemens Molecular Imaging, Inc., Knoxville, TN) as previously reported.^22^ To ensure accurate and unbiased results, an inter-scanner normalization method was implemented, and images were corrected for scatter and motion.^23^ [^18^F]-FEOBV was prepared following the methodology described previously.^24^ [^18^F]-FEOBV delayed dynamic imaging was performed over 30 minutes (in six 5-minute frames) starting 3 hours after an intravenous bolus dose injection of 8 mCi [^18^F]-FEOBV.^25^ We co-registered MRI to PET images of each subject using MRI-PET registration statistical parametric mapping (SPM) software (SPM12; Welcome Trust Centre for Neuroimaging, University College, London, England [https://www.fil.ion.ucl.ac.uk/spm/software/spm12/]). [^18^F]-FEOBV PET images were analyzed non-invasively using a supratentorial white matter reference tissue approach as previously reported.^26,27^ Distribution volume ratios (DVR) were calculated from ratios of the averaged six delayed imaging PET frames 3 hours post injection and white matter reference tissues.^27^ The reference region (supratentorial white matter) was defined by excluding voxels below the ventricles and near cortical areas, followed by erosion. Parametric images were then generated by dividing voxel values in the respective activity images by the mean activity in this reference region. A Muller-Gartner partial-volume correction method was used to remove the partial volume effect (PVE) on the parametric PET images. Individual subject PVE corrected parametric [^18^F]-FEOBV studies were normalized to the study-specific template in Montreal Neurological Institute (MNI) space using high-dimensional DARTEL registration.

### Participant Classification and Longitudinal Cognitive Assessments

Participants were categorized based on longitudinal cognitive domain changes using the MDS PD-MCI level II criteria; greater than 1.5 SD decline on at least two neuropsychological tests in one cognitive domain or a greater that 1.5 SD decline in single tests within two cognitive domains.^28^ Classification was based on cognitive trajectories between baseline and 2-year follow-up visits. There were 4 classes: 1) persistent normal, not meeting criteria for MCI at baseline or at two-year follow-up; 2) persistent MCI, meeting MCI criteria at both baseline and at 2-year follow-up visits; 3) MCI reversion, meeting criteria for MCI at baseline but not at 2-year follow up visits; 4) MCI conversion, not meeting criteria for MCI at baseline but meeting criteria for MCI at 2-year follow up visits. To further investigate longitudinal cognitive changes over the 2-year interval between baseline and follow-up visits, percent changes in z-scores (z-score baseline - z-score follow up/z-score baseline x 100) were computed to investigate whether regional patterns of [^18^F]-FEOBV binding were associated with longitudinal cognitive changes. Negative percent z-score changes represented decline and positive percent z-score changes represented improvement or stability from baseline.

### Data and Statistical Analysis

Our primary objective was to assess whether baseline brain regional [^18^F]-FEOBV uptake is correlated with longitudinal cognitive changes in a group of cognitively heterogeneous PD participants. We employed a four-step analytic approach. First, we explored the differences in baseline [^18^F]-FEOBV binding at the voxel-level by comparing the 4 clinically defined classes - persistent normal, persistent MCI, reverting MCI, and converting MCI, - with HC. Second, we explored associations between baseline [^18^F]-FEOBV binding and global composite percent z-score changes at the voxel level (voxel-wise SPM12 correlation analyses). Third, we utilized a pattern identified as significant from the voxel-based analysis (see below) to study associations between baseline regional [^18^F]-FEOBV binding and cognitive changes as assessed by global composite percent z-score changes and percent z-score changes for individual cognitive domains (vision, language, memory, executive and attentive) with Volume of Interest (VOI) linear regression analyses. Finally, we utilized the pattern derived from the voxel-based analysis to determine if there were significant baseline differences between the four distinct classes utilizing a between groups ANOVA.

1) Voxel-wise SPM12 analyses of clinically defined classes: A voxel-based two-sample t-test within SPM12 was utilized to compare HC [^18^F]-FEOBV PET and baseline [^18^F]-FEOBV PET for each of the 4 clinically defined classes. For the peak cluster analysis, significance thresholds after false discovery (FDR) correction and family wise error (FWE) correction were set at P < 0.05. The statistical threshold was *p* < 0.01, cluster extent (K)>100.
2) Voxel-wise SPM12 correlation analyses (exploratory approach): We employed a voxel-wise SPM12 regression model, entering global composite percent z-score change as the primary variable of interest. We corrected for baseline cognitive status using the baseline composite z-scores as a covariate of no interest. For the peak cluster analysis, significance thresholds after false discovery (FDR) correction and family wise error (FWE) correction were set at *p* < 0.05, cluster extent (K) >100.
3) Cluster VOI-based Regression Analyses Using Significant Voxel Volume of Baseline [^18^F]-FEOBV Clusters Associating with Interval Cognitive Changes (VOI-based approach): To further investigate the association between baseline regional [^18^F]-FEOBV binding and cognitive status at follow-up, we utilized significant clusters from voxel-wise SPM12 correlation analyses (step 2 results) as VOIs to predict cognitive outcomes at FU using linear regression models. Specifically, we employed the REX toolbox in Matlab to extract DVR mean values from these cluster VOIs in participants’ [^18^F]-FEOBV scans. These extracted values served as regressors in linear regression models to explore their association with cognitive status at follow-up. This included examining the global composite z-score and z-scores for individual cognitive domains (memory, vision, executive function, language, and attention) for each patient. Distributions of residuals were examined to ensure assumptions of the model were not violated utilizing Kolmogorov-Smirnov test of normality. In cases of violation, the dependent variable was transformed with rank values to reduce the skewness and the probability of having abnormally distributed results. These statistical analyses were run with SPSS (version 29.0).
4) Between Groups ANOVA: To determine baseline regional [^18^F]-FEOBV binding differences in the cluster uptake values (step 2 results) between the 4 clinically defined classes. These statistical analyses were run with SPSS (version 29.0).

### Data availability

Study data are available from the corresponding author upon reasonable request.

## RESULTS

### Participant Clinical Features

This longitudinal study involved 75 subjects with PD (54 males; 21 females). Baseline and 2-year follow-up visit data are summarized in **Table 1**. Paired samples t-tests revealed significant increases in LED, MDS-UPDRS Part III, and vision z-scores between baseline and follow-up visits. Based on MDS PD-MCI level II criteria, 41 (54.6%) PD participants were classified as persistent normal; 21 (28.00%) as persistent MCI; 7 (9.33%) as reverted MCI; and 6 (8.00%) as converted MCI. The persistent normal group had an average global composite z-score of −0.07 (0.35) at baseline and an average global composite z-score of 0.17 (0.30) at follow up. The persistent MCI group had an average global composite z-score of −1.102 (0.58) at baseline and an average global composite z-score of −1.12 (0.61) at follow up. The reverted MCI group had an average baseline global composite z-score of −0.28 (0.23) and a follow-up average global z-score of −0.03 (0.29). The converted MCI group had a baseline average global composite z-score of - 0.22 (0.25) and a follow-up average global z-score of −0.62 (0.37). We conducted a one-way ANOVA to determine any significant demographic differences between the four distinct classes. A Tukey post-hoc test revealed at baseline there were significant differences between MDS UPDRS-III scores (F(3) = 3.009, *p* = .027), age (F(3) =4.615, *p* =.004), and education level (F(3) = 4.008, *p* =.017), between the persistent normal and persistent MCI groups. See **Supplementary Table 2** for more comprehensive group demographics and clinical features.

**Table 1:** Demographic, Clinical and Cognitive Data for Baseline and Follow-up Visits.

| PD | Baseline | Follow-Up | Two-sided $p^1$ |
| --- | --- | --- | --- |
| Age | 66.93 (6.67)<sup>2</sup> | 68.88 (6.68) | **0.001** |
| Gender | Females: 21<br>Males: 54 | NA | NA |
| Education (years) | 16.16 (2.51) | NA | NA |
| Disease Duration (years) | 5.60 (3.98) | NA | NA |
| Hoehn & Yahr stage | 2.31 (0.59) | 2.41(0.55) | 0.176 |
| MDS-UPDRS Part III | 31.55 (11.73) | 37.75 (13.48) | **0.001** |
| LED (mg) | 593.28 (353.18) | 730.78 (464.96) | **0.002** |
| MoCA | 26.99 (2.37) | 27.04 (2.38) | 0.834 |
| Memory z-scores | -0.19 (0.81) | -0.17 (0.95) | 0.816 |
| Language z-scores | -0.27 (0.82) | -0.31 (0.77) | 0.601 |
| Attention z-scores | -0.46 (0.89) | -0.47 (0.90) | 0.763 |
| Executive z scores | -0.39 (1.01) | -0.34 (1.01) | 0.351 |
| Vision z-scores | -0.25 (0.74) | -0.04 (0.66) | **0.004** |
| Global z-scores | -0.31 (0.65) | -0.27 (0.70) | 0.210 |

*Abbreviations: MDS-UPDRS, Movement Disorder Society–Unified Parkinson’s Disease Rating Scale; LED, levodopa equivalent dose.*
<sup>1</sup>Paired samples t-tests
<sup>2</sup>Means (standard deviations) are presented for numerical variables.

### Voxel-Wise Group Comparisons: Cognitive Classes vs HC

#### Persistent Normal

The whole brain voxel-based analysis (FDR and FWE corrected at the cluster level, *P <* 0.05) revealed significant differences between regional [^18^F]-FEOBV binding between HC and the persistent normal group. The persistent normal group displayed higher [^18^F]-FEOBV binding than HC in the anterior cingulate, frontal medial orbital cortices, para-hippocampus, hippocampus, right insula, putamen, and right caudate. This group also showed reduced [^18^F]-FEOBV binding compared HC in the cuneus, precuneus, paracentral lobe, posterior temporal cortices, occipital cortices and posterior cingulum (**Figure 1A**).

**Figure 1:**
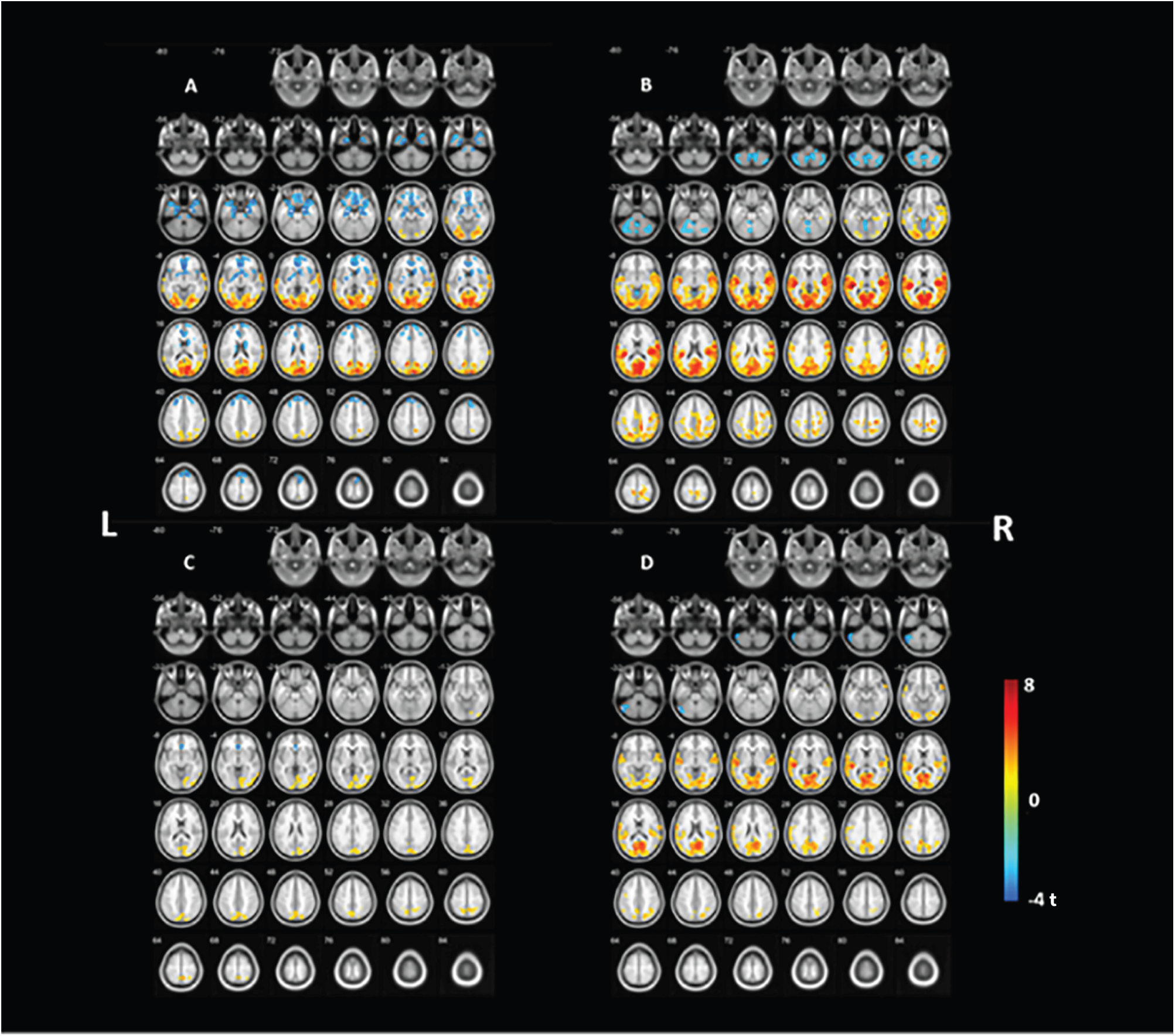
Voxel-Wise Group Comparisons: Cognitive Classes vs Healthy Controls. Statistical parametric voxel-based analysis of differences in baseline [^18^F]-FEOBV binding of 4 cognitive PD classes and healthy controls (HC); (A) Persistent Normal, (B) Persistent MCI, (C) Reverted MCI, and (D) Converted MCI. Blue denotes increased [^18^F]-FEOBV binding in PD versus HC. Red denotes decreased [^18^F]-FEOBV binding in PD versus HC. Thresholds set at *p* < 0.01, K>100 in all voxel-wise comparisons.

#### Persistent MCI

Whole brain voxel-based analysis (FDR and FWE corrected at the cluster level, *P <* 0.05) revealed significant differences between regional [^18^F]-FEOBV binding between HC and the persistent MCI group. Increased [^18^F]-FEOBV binding in the persistent MCI group compared to HC was identified primarily in the cerebellum, and supplementary motor area. Reduced [^18^F]-FEOBV binding compared to HC was seen primarily in the right more than left para-hippocampus, thalamus, hippocampus, supramarginal cortices right more than left, posterior temporal cortices, parietal cortices, occipital cortices, cuneus, and posterior cingulate. (**Figure 1B**).

#### Reverted MCI

The reverted MCI group showed a trend toward increased [^18^F]-FEOBV binding compared to HC in the anterior cingulate and left caudate, left thalamus, and left insula (uncorrected *p <* 0.05). Reduced [^18^F]-FEOBV binding in the reverted MCI group compared to the HC was also identified in the precuneus, cuneus, occipital cortices and the posterior cingulate (FDR and FWE corrected at the cluster level, *P <* 0.05) (**Figure 1C**).

#### Converted MCI

The converted MCI group displayed a trend toward higher [^18^F]-FEOBV binding than HC in the left cerebellum (uncorrected *p <* 0.05). Reduced [^18^F]-FEOBV binding in the converted MCI group compared to the HC was identified primarily in the posterior temporal cortices, parietal cortices, occipital cortices, cuneus, and posterior cingulate (FDR and FWE corrected at cluster level, *p <* 0.05). (**Figure 1D**).

Mean percent difference voxel [^18^F]-FEOBV DVR from controls and significant cluster extent across cognitive classes is shown in **Table 2**.

**Table 2:** Mean Percent Difference Voxel [^18^F]-FEOBV DVR and Cluster Extent Across Cognitive Classes.

| Group | Hypo<br>Difference<br>(percent) | Cluster Extent | Hyper<br>Difference<br>(percent) | Cluster Extent |
| --- | --- | --- | --- | --- |
| Persistent<br>Normal | -15.2% | 22,074 | 12.8% | 13,346 |
| Persistent<br>MCI | -20.4% | 48,338 | 17.8% | 3,802 |
| MCI<br>Reversion | -24.0% | 4,769 | 25.7% | 252 |
| MCI<br>Conversion | -27.1% | 21,162 | 34.8% | 840 |

### Voxel-Based Whole Brain [^18^F]-FEOBV Binding Correlates of Global Cognitive Changes

We found significant correlations between global cognitive changes, measured as global composite percent z-score changes, and baseline regional [^18^F]-FEOBV binding. Lower [^18^F]-FEOBV binding in several regions was associated with cognitive decline. Significant correlations (cluster-FDR and FWE corrected significant at *p* <0.05) were seen primarily in mid to posterior cingulum, precuneus, parietal-lateral occipital, right more the left superior temporal, opercular, right more than left precentral and postcentral gyrus, right para-hippocampus, and right insula. This constellation was collectively labeled as the posterior, limbic and paralimbic (PLP) cluster (**Figure 2**).

**Figure 2:**
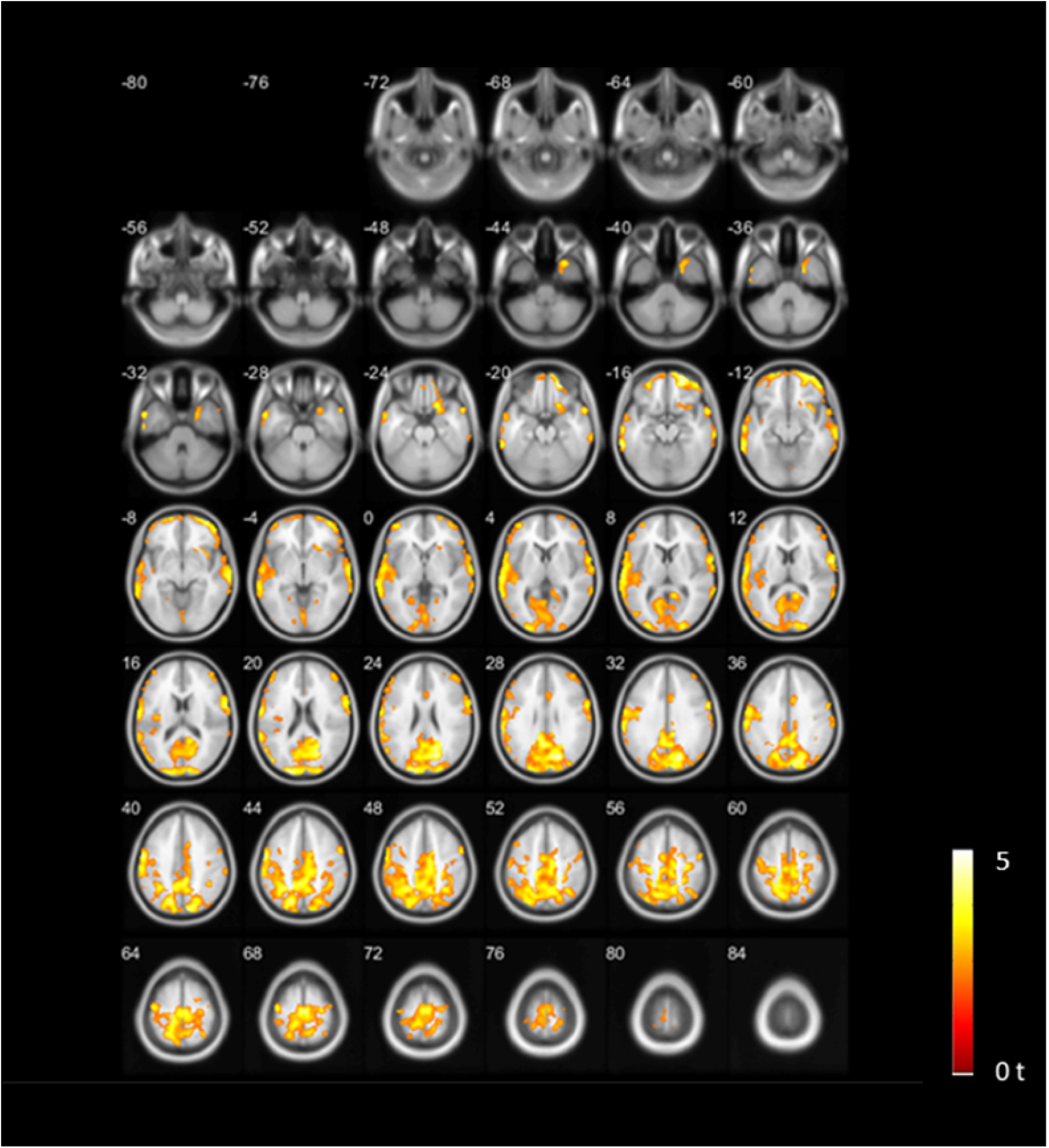
Voxel-Based Regional [^18^F]-FEOBV Binding Correlates of Global Cognitive Changes. Voxel-based analysis of correlations between baseline [^18^F]-FEOBV binding and cognitive changes as assessed by percent change in global composite z-scores. Significant correlations (cluster-FDR corrected significant at *p* < 0.05) were seen primarily in mid to posterior cingulum, precuneus, parietal-lateral occipital, right more the left superior temporal, opercular, right more than left precentral and postcentral gyrus, right parahippocampus, and right insula. This constellation was collectively labeled as the posterior, limbic and paralimbic (PLP) cluster. Threshold was set at *p* < 0.05, K > 100, and cluster level significant with FDR *p* < 0.05, controlling for baseline cognition.

### VOI-Based Analyses

#### VOI baseline [^18^F]-FEOBV Clusters Associated with Interval Global Cognitive Changes

We observed that lower mean baseline PLP cluster [^18^F]-FEOBV binding was significantly associated with lower z-scores in global cognition, visual, memory, language, attention, and executive cognitive domains at two-year follow-up after controlling for disease duration (**Table 3**). Baseline PLP cluster [^18^F]-FEOBV binding exhibited stronger associations with executive and attentional z-scores at follow-up, explaining 25% and 26% of their variance, respectively. For the other domains, the variance explained was lower, ranging from 6% to 11% (**Table 3**).

**Table 3:** Results of Linear Regression Analyses Between Baseline Posterior, Limbic and Paralimbic (PLP) Cluster [^18^F]-FEOBV binding and Follow-up (FU) Cognitive Z-Scores.

| Model | Sum of Squares | Adjusted R<sup>2</sup> | $\beta$ | t-value | F | *p* | 95% CI | |
| --- | --- | --- | --- | --- | --- | --- | --- | --- |
|  |  |  |  |  |  |  | Lower | Upper |
| Global z-score FU | 9.447 | 0.239 | 4.026 | 4.758 | 12.594 | 0.001 | 2.339 | 5.713 |
| Vision z-score FU | 2.762 | 0.060 | 2.236 | 2.528 | 3.371 | 0.040 | 0.473 | 3.999 |
| Language z-score FU | 5.271 | 0.096 | 3.170 | 3.133 | 4.914 | 0.010 | 1.153 | 5.188 |
| Memory z-score FU | 8.679 | 0.107 | 3.703 | 3.000 | 5.437 | 0.004 | 1.242 | 6.164 |
| Executive z-score FU | 20.290 | 0.247 | 5.590 | 4.600 | 12.118 | 0.001 | 3.168 | 8.013 |
| Attention z-score FU | 16.956 | 0.263 | 5.431 | 5.084 | 14.183 | 0.001 | 3.301 | 7.561 |

#### Comparison of Baseline Posterior, Limbic and Paralimbic Cluster [^18^F]-FEOBV Binding Among Cognitive Classes

One-way ANOVA revealed that there was a statistically significant difference in baseline PLP uptake cluster [^18^F]-FEOBV binding between groups (F(3), = 3.252, p = .027). A Tukey post-hoc test revealed significantly higher uptake value in the mean PLP cluster [^18^F]-FEOBV binding between the persistent normal group and the persistent MCI group (*p* = 0.035, 95% C.I. = [0.003, 0.117]). There were no significant differences between the persistent normal and MCI converted groups (*p* = 0.204) or between the persistent normal and reverted MCI groups (*p* =.852) (**Table 4**).

**Table 4:** Descriptive Statistics for Baseline Posterior, Limbic and Paralimbic (PLP) Cluster [^18^F]-FEOBV binding Among the Four Cognitive Classes.

| Group | N | Mean Voxel<br>DVR | Std Deviation | 95% Confidence Interval Mean |  |
| --- | --- | --- | --- | --- | --- |
|  |  |  |  | Lower | Upper |
| Persistent Normal | 41 | 0.60 | 0.07 | 0.58 | 0.63 |
| Persistent MCI | 21 | 0.54 | 0.02 | 0.51 | 0.58 |
| MCI Reversion | 7 | 0.57 | 0.11 | 0.48 | 0.67 |
| MCI Conversion | 6 | 0.53 | 0.118 | 0.41 | 0.65 |

## DISCUSSION

This study aimed to investigate the relationship between baseline regional [^18^F]-FEOBV binding in a heterogeneous group of mild-moderate, non-demented PD subjects and cognitive changes over a two-year period. After classifying our subject population on the basis of cognitive stability or change in normal versus PD-MCI status, we found distinct patterns of baseline [^18^F]-FEOBV binding in the four categorized groups relative to a control group. These differing patterns of regional [^18^F]-FEOBV binding included both regions with diminished and increased [^18^F]-FEOBV binding. Using a voxel-based analysis, we found an association between cognitive decline, assessed by percent change in global composite z-score changes, and a constellation of regional [^18^F]-FEOBV binding deficits. The voxel-based analysis results were used to derive a group of regions – the PLP [^18^F]-FEOBV cluster – for VOI analysis. Linear regression analyses using the PLP [^18^F]-FEOBV binding values as regressor demonstrated that baseline PLP [^18^F]-FEOBV binding decreases predicted global cognitive changes and domain-specific changes at follow-up. Direct comparison of baseline regional PLP [^18^F]-FEOBV binding among the four defined groups via ANOVA confirmed significant differences between the persistent normal and persistent MCI groups.

[^18^F]-FEOBV is a high affinity, specific ligand for VAChT, a protein uniquely expressed by cholinergic neurons. [^18^F]-FEOBV PET emerged as an anatomically resolute measure of regional cholinergic terminal density, with strong correlation between regional distribution of [^18^F]-FEOBV binding *in vivo* and histological studies of human cholinergic systems anatomy.^29,30^ Reductions in regional brain [^18^F]-FEOBV binding are parsimoniously explained by loss of cholinergic terminals. For cortical regions, this inference is supported by correlations between regional cortical [^18^F]-FEOBV binding and atrophy of basal forebrain nuclei originating cholinergic projection neurons.^26,31,32^ The most plausible explanation for elevated regional [^18^F]-FEOBV binding is upregulation of VAChT expression, likely reflecting increased cholinergic neurotransmission. The gene encoding VAChT, *SLC18A3*, is embedded within the first intron of the gene encoding choline acetyltransferase (*CHAT*) and there appears to be coordinate regulation of expression of these two gene products.^33^ Increased VAChT expression implies increased CHAT expression and increased synthetic demand for acetylcholine. ChAT activity is a conventional measure of the magnitude of cholinergic neurotransmission. In post-mortem studies of MCI and mild AD, for example, regional ChAT activity increases were inferred to reflect increased cholinergic neurotransmission.^34^ Alternative, less likely, explanations for increased [^18^F]-FEOBV binding are increased cholinergic terminal density due to increased absolute numbers of terminals or marked regional atrophy with relative sparing of cholinergic terminals.^19^ Increased cholinergic neurotransmission in the setting of neurodegenerative disorders may reflect a form of compensation as cholinergic systems become more active when other brain systems fail.^34,35^

This is the first longitudinal analysis of the relationship between cognitive changes and brain cholinergic systems integrity using anatomically resolute brain [^18^F]-FEOBV PET imaging. Across the four defined subject groups, baseline cholinergic denervation was observed in the occipital, parietal, and temporal cortices compared to HC with the most marked regional denervation occurring in the persistent MCI group. These results are consistent with prior studies using [^18^F]-FEOBV or AChase PET. PD subjects with persistent normal cognition, however, exhibited increased [^18^F]-FEOBV binding in the anterior cingulate, frontal medial orbital cortices, para-limbic, limbic, and anterior cingulum regions. Similarly, the reverted MCI group displayed increased [^18^F]-FEOBV binding in the anterior cingulate and limbic regions. These findings suggest a similar regional pattern of increased, perhaps compensatory, cholinergic neurotransmission in the persistent normal and reverting MCI groups. Increased [^18^F]-FEOBV binding was also seen in some striatal regions. Overlapping regional [^18^F]-FEOBV binding patterns were also seen in the persistent MCI and converting MCI groups. Both the persistent MCI and converting MCI groups exhibited diminished [^18^F]-FEOBV binding in several cortical regions with no forebrain regions exhibiting increased regional [^18^F]-FEOBV binding. Both the persistent MCI and converting MCI group exhibited increased cerebellar [^18^F]-FEOBV binding, more extensive in the persistent MCI group than the converting MCI group. Non-significant trends toward spatially more limited higher [^18^F]-FEOBV binding were also observed for the reverted and converted MCI subgroups.

Assuming that increased [^18^F]-FEOBV binding indicates increased cholinergic neurotransmission, the observed increases in [^18^F]-FEOBV binding in different brain regions suggest compensatory activity in several cholinergic systems. Anterior cingulate, orbitofrontal cortices, and parahippocampal region [^18^F]-FEOBV binding increases in the persistent normal group suggest compensatory processes involving cholinergic projections originating in the basal forebrain complex.^36^ Striatal region [^18^F]-FEOBV binding increases suggest increased compensatory striatal cholinergic interneuron activity with possible additional contributions from the pedunculopontine nucleas and lateral dorsal tegmentum nuclei.^37^ Both the persistent MCI and converting groups exhibited increased [^18^F]-FEOBV binding in the cerebellum, consistent with increased activity of cholinergic medial vestibular complex cholinergic neurons. Our results are broadly consistent with some prior studies. van der Zee et al. used [^18^F]-FEOBV PET to study a group of *de novo* PD subjects, some with normal cognition and some classified as PD-MCI, demonstrating both reductions and increases of regional [^18^F]-FEOBV binding.^17^ These results are not identical to our findings, but there is significant overlap with both cognitively normal and PD-MCI subjects exhibiting diminished [^18^F]-FEOBV binding in posterior cortical regions and cognitively normal PD subjects exhibiting increased [^18^F]-FEOBV binding in some frontal regions and the cerebellum. In a small study of isolated REM sleep behavior disorder subjects later converting to Multiple System Atrophy, Bedard et al. reported increased regional [^18^F]-FEOBV binding.^38^

[^18^F]-FEOBV binding decreases in a number of more posterior cortical regions is consistent with results of several prior cross-sectional studies. A potential explanation for the complex pattern of increased [^18^F]-FEOBV binding in more anterior-midline cortical regions and reduced more posterior cortical [^18^F]-FEOBV binding may lie in the biology of basal forebrain corticopetal cholinergic projections. Chakraborty et al. presented data indicating that basal forebrain projections to more midline cingulo-insular cortices exhibit more diffuse axonal arboratization than those to more distant unimodal cortices.^39^ Providing metabolic support for the more distal terminals may be particularly demanding and they may be particularly prone to degeneration as basal forebrain cholinergic neurons attempt to compensate for other failing systems. Regardless, cumulative results suggest that changes in cholinergic systems function and integrity are dynamic across the range of PD and stress the importance of longitudinal analyses.

Our results also strongly suggest that varying cholinergic systems changes are significant contributors to the heterogeneity of cognitive changes in PD. We demonstrated an association between baseline [^18^F]-FEOBV PET results and subsequent cognitive changes over a two year period. Lower baseline [^18^F]-FEOBV binding in posterior occipital, temporal, and parietal cortices, and limbic regions exhibited correlations with cognitive decline across this heterogeneous sample. Notably, [^18^F]-FEOBV binding in the pre-defined PLP cluster emerged as a predictor of cognition at two-year follow-up, both globally and spanning various cognitive domains, with baseline PLP deficits exhibiting the strongest relationship to variance within the attention and executive function domains. This aspect of our results is consistent with known functions of forebrain cholinergic systems.

The baseline cholinergic deficits associated with cognitive decline may also be useful predictors of future cognitive status. Mean [^18^F]-FEOBV binding in the PLP cluster effectively discriminated between the persistent normal and persistent MCI groups, consistent with PLP [^18^F]-FEOBV binding predicting longitudinal cognitive changes. Our findings of baseline [^18^F]-FEOBV PET imaging potentially predicting cognitive changes over a two-year period highlights the potential use of neuroimaging techniques for prediction of cognitive decline. Longitudinal MRI studies demonstrated that atrophy of the basal forebrain complex and Nucleus basalis of Meynert predict cognitive changes in PD.^40,41^

This study has limitations. We enrolled subjects with mild to moderate PD and the 2-year follow-up spans only a subset of the clinical course of PD. The [^18^F]-FEOBV upregulation findings for the converting and reverting MCI groups failed to survive multiple comparisons when compared to HC. These negative results may be due to small sample sizes (n=7 in reverting MCI and n=6 in converting MCI). The reverting MCI group may be composed of a more heterogeneous sample with various confounding factors contributing to reversion, including medication changes, mood disorder changes, or cognitive fluctuations. Though we incorporated disease duration into our analysis, there were some baseline clinical differences between the persistent normal and persistent MCI groups in education level, age, and MDS-UPDRS_III_. These findings are not unexpected as higher motor severity has been correlated with cognitive impairment in PD.^42^

Our findings reinforce the concept that cholinergic systems changes are important actors in cognitive changes in PD. The four classes of cognitive changes over a two-year period had distinctive bidirectional cholinergic regional cholinergic terminal density distributions. We found a distinctive pattern of cholinergic vulnerability associated with subsequent two-year cognitive decline. Varying cholinergic systems changes are likely contributors to the heterogeneous character of cognitive decline in PD. Cholinergic imaging or related biomarkers may be useful tools for predicting the course of cognitive changes in PD, for stratification in clinical trials, and may also be useful in defining different subtypes of PD. These results may identify useful targets for interventions aimed at ameliorating cognitive dysfunctions in PD. Expanded longitudinal studies correlating the relationships between brain cholinergic systems changes and cognitive changes in PD are warranted.

## ACKNOWLEDGMENTS

We thank our research participants and the staff of the University of Michigan PET Center. Supported by P50NS123067, R21NS144749, P50NS091856, R01AG073100, I01RX003397, I01 RX001631 and the Parkinson’s Foundation.

## Author Roles

1) Conception and design of the study: TB, PK, GC, NB, RA

2) Acquisition and analysis of data: TB, PK, GC

3) Drafting a significant portion of the manuscript: TB, PK, GC, JB, RA, NB

## Conflicts of Interest

RLA serves on the Data Safety and Monitoring boards for the CELIA, SIGNAL-AD, TOPAS-MSA, and Zilgenersen trials. The other authors report no conflicts of interest.

---

**Supplementary Table 1:** Comprehensive Neuropsychological Test Battery and Domain Specific Categorization.

| **Domain** | **Neuropsychological Tests** |
| --- | --- |
| **Memory** | California verbal learning test: immediate recall |
|  | California verbal learning test: delayed free recall |
|  | Wechsler Memory Scale |
| **Attention** | Stroop 2: color test |
|  | Delis–Kaplan Executive Function System, Trail Making Test 2: number sequencing |
|  | Symbol Digit Modalities Test |
|  | Wechsler Adult Intelligence Scale: digit span backward |
| **Executive function** | Stroop 4: Adjusted Color Word Test |
|  | Delis–Kaplan Executive Function System, Trail Making Test 4: letter–number sequencing |
|  | Wechsler Adult Intelligence Scale: matrix reasoning |
|  | Letter fluency |
| **Language** | Boston Naming Test |
|  | Verbal fluency: animals |
| **Visual function** | Judgment of line orientation |
|  | Parkinson’s Disease–Cognitive Rating Scale: Clock Test |

**Supplementary Table 2:**
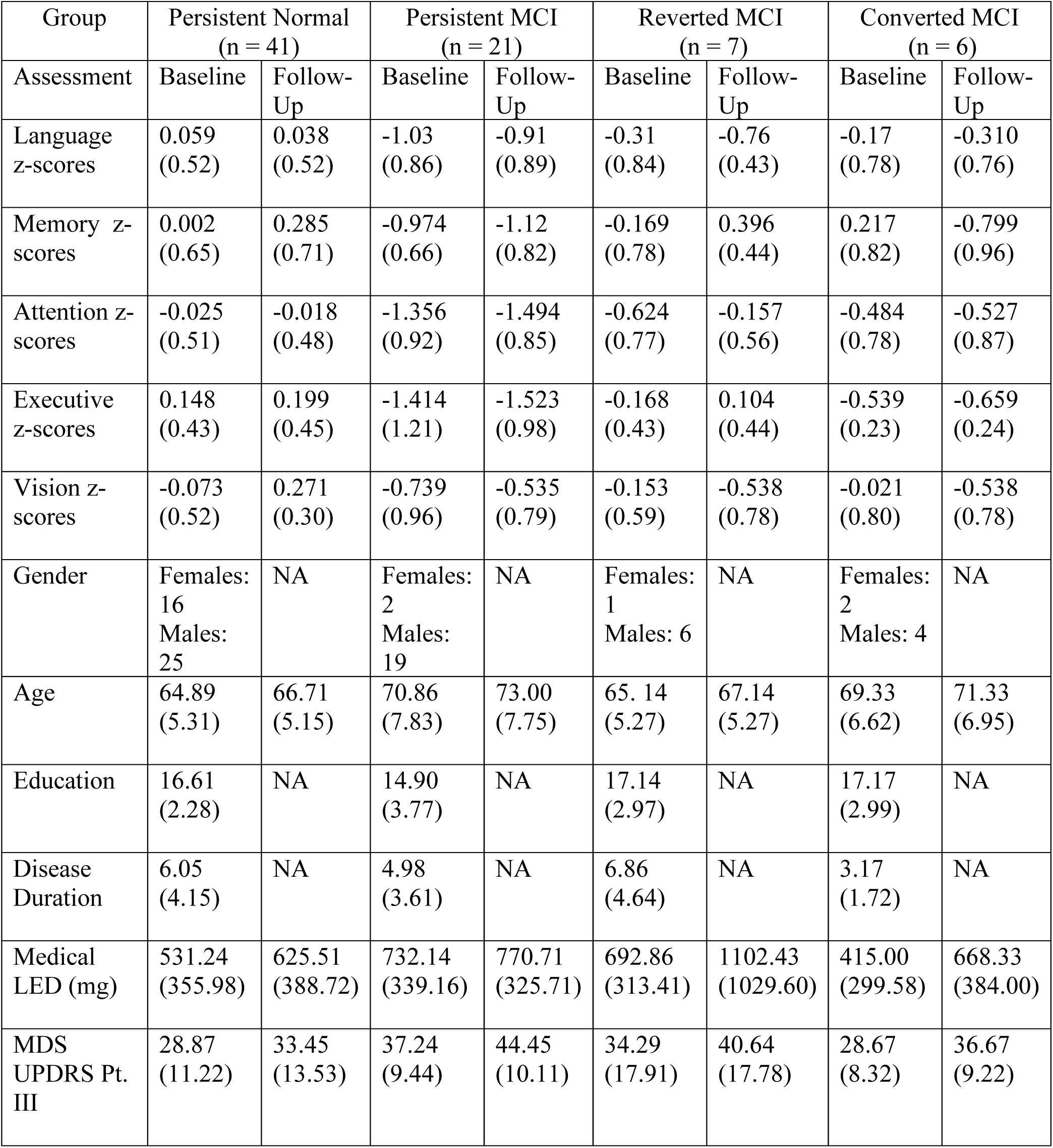
Descriptive Statistics for Longitudinal Changes Between Cognitive Classes.

| Group | Persistent Normal<br>(n = 41) |  | Persistent MCI<br>(n = 21) |  | Reverted MCI<br>(n = 7) |  | Converted MCI<br>(n = 6) |  |
| --- | --- | --- | --- | --- | --- | --- | --- | --- |
| Assessment | Baseline | Follow-Up | Baseline | Follow-Up | Baseline | Follow-Up | Baseline | Follow-Up |
| Language z-scores | 0.059<br>(0.52) | 0.038<br>(0.52) | -1.03<br>(0.86) | -0.91<br>(0.89) | -0.31<br>(0.84) | -0.76<br>(0.43) | -0.17<br>(0.78) | -0.310<br>(0.76) |
| Memory z-scores | 0.002<br>(0.65) | 0.285<br>(0.71) | -0.974<br>(0.66) | -1.12<br>(0.82) | -0.169<br>(0.78) | 0.396<br>(0.44) | 0.217<br>(0.82) | -0.799<br>(0.96) |
| Attention z-scores | -0.025<br>(0.51) | -0.018<br>(0.48) | -1.356<br>(0.92) | -1.494<br>(0.85) | -0.624<br>(0.77) | -0.157<br>(0.56) | -0.484<br>(0.78) | -0.527<br>(0.87) |
| Executive z-scores | 0.148<br>(0.43) | 0.199<br>(0.45) | -1.414<br>(1.21) | -1.523<br>(0.98) | -0.168<br>(0.43) | 0.104<br>(0.44) | -0.539<br>(0.23) | -0.659<br>(0.24) |
| Vision z-scores | -0.073<br>(0.52) | 0.271<br>(0.30) | -0.739<br>(0.96) | -0.535<br>(0.79) | -0.153<br>(0.59) | -0.538<br>(0.78) | -0.021<br>(0.80) | -0.538<br>(0.78) |
| Gender | Females: 16<br>Males: 25 | NA | Females: 2<br>Males: 19 | NA | Females: 1<br>Males: 6 | NA | Females: 2<br>Males: 4 | NA |
| Age | 64.89<br>(5.31) | 66.71<br>(5.15) | 70.86<br>(7.83) | 73.00<br>(7.75) | 65.14<br>(5.27) | 67.14<br>(5.27) | 69.33<br>(6.62) | 71.33<br>(6.95) |
| Education | 16.61<br>(2.28) | NA | 14.90<br>(3.77) | NA | 17.14<br>(2.97) | NA | 17.17<br>(2.99) | NA |
| Disease Duration | 6.05<br>(4.15) | NA | 4.98<br>(3.61) | NA | 6.86<br>(4.64) | NA | 3.17<br>(1.72) | NA |
| Medical LED (mg) | 531.24<br>(355.98) | 625.51<br>(388.72) | 732.14<br>(339.16) | 770.71<br>(325.71) | 692.86<br>(313.41) | 1102.43<br>(1029.60) | 415.00<br>(299.58) | 668.33<br>(384.00) |
| MDS UPDRS Pt. III | 28.87<br>(11.22) | 33.45<br>(13.53) | 37.24<br>(9.44) | 44.45<br>(10.11) | 34.29<br>(17.91) | 40.64<br>(17.78) | 28.67<br>(8.32) | 36.67<br>(9.22) |

## Notes

### Competing Interest Statement

Roger Albin serves on the Data Safety and Monitoring Boards for the CELIA, SIGNAL-AD, TOPAS-MSA, and Zilgenersen trials. The other authors report no competing interests.

### Funding Statement

This study was funded by the National Institutes of Health, the Department of Veterans Affairs, and the Parkinsons Foundation.

### Author Declarations

The IRBs of the University of Michigan and the VA Ann Arbor Health System gave ethical approvals for this work.

